# Immediate and Early Engagement of Same-Day Antiretroviral Therapy Initiation among newly diagnosed people living with HIV in Urban Zambia: A Retrospective Cohort Study

**DOI:** 10.1101/2022.08.25.22278881

**Authors:** Mpande Mukumbwa-Mwenechanya, Obvious Nchimunya Chilyabanyama, Estella Kalunkumya, Violet Kunda, Kombatende Sikombe, Jake M Pry, Godwin Nyirenda, Mwansa Lumpa, Anjali Sharma, Samuel Bosomprah, Carolyn Bolton-Moore

## Abstract

**Introduction:** As Zambia moves towards attaining human immunodeficiency virus (HIV) epidemic control, it is clear significant efforts are required to facilitate achievement of UNAIDS treatment targets by 2030. To accelerate progress towards global target of 95% of people living with HIV (PLHIV) knowing their status, country is promoting community based HIV testing and same day antiretroviral therapy (ART) initiation. However, there are uncertainties around acceptability of this strategy and how it affects immediate and early engagement in program settings.

**Methods:** We included all newly diagnosed PLHIV aged 18 years or older and provided same day ART initiation between October 2018 and January 2019 in Lusaka District. Immediate engagement was estimated as proportion of newly diagnosed PLHIV who visited the health facility at least once within 14 days after same-day ART initiation, whereas early engagement as proportion of newly diagnosed PLHIV active 6 months after same day ART initiation. Pearson’s chi-squared test was used to assess association of outcomes with key background characteristics.

**Results:** Of 12,777 newly diagnosed PLHIV who initiated same day ART 7,943 (62%) were tested and initiated in the community. Overall, 6,257 (49%) engaged within 14 days (median 15, IQR:13 37). Older individuals (36-49 years) were more likely to be engaged at 14 days (aRR 1.29: 95%CI 1.06-1.18; p<0.001) and retained at 6 months (aRR:1.27;95%CI 1.21-1.34P<0.001) whilst risk of attrition at 6 months was highest in younger ages (18-24 years) (aRR 0.79;95 %CI 0.76-0.82; p<0.001).

**Conclusion:** To adequately address the HIV epidemic targeted engagement approaches are required particularly in the younger ages.

## Introduction

Human immunodeficiency virus (HIV) epidemic control remains a public health concern, despite substantial progress made in the last two decades. The race to achieve HIV/AIDS elimination by 2030 has been set as the UNAIDS 95-95-95 global targets (i.e. Ninety-five percent of all people living with HIV (PLHIV) know their HIV status, 95% of all people with diagnosed HIV infection receive sustained antiretroviral therapy (ART), and 95% of all people receiving ART have viral suppression).^1^ Zambia is striving to achieve HIV epidemic control, and efforts have contributed to lowering HIV prevalence however, current estimates of 71-88-89 among 15-59-year-old PLHIV fall short of satisfying the global target of 95-95-95 by 2030.^2^ The gaps in attaining these treatment targets, particularly the first and second 95, underscore the need for intensive efforts towards attaining UNAIDS 2030 treatment targets.^3^

The Zambian Government has mandated universal HIV testing services (HTS), which provides an opportunity to screen all individuals for HIV and immediately treat all HIV-infected individuals regardless of cluster of differentiation 4 (CD4) count or world health organisation (WHO) clinical stage. Whilst primarily conducted at health facilities HTS also occurs outside the health facility in the “community” to expand individuals’ knowledge of their HIV status, accelerate linkage to care, improve ART initiation, limit the spread of infection, reach priority and key vulnerable populations (KVPs) and consequently ensure that no one is left behind in the fight against HIV.^2^ The HTS package provides a full range of services including pre-and post-test counselling, linkage to appropriate HIV prevention, treatment, and care services; and referral to other appropriate clinical and support services. Additionally, individuals who test positive are initiated on ART and provided with antiretroviral medication for 14 days in which the newly initiated PLHIV must visit a health facility within 14 days of initiation to collect a 1-month supply and thereafter 3-month supply.

Outside health facility “Community-based” as well as facility-based strategies of rapid ART initiation are being utilized to test as many people as possible so as to expand the HIV status knowledge, address the poor linkage of care and ensure that no one is left behind.^4-9^ Whilst the advantages of immediate and sustained ART treatment are well established, the benefits do not fully materialize due to delayed and intermittent engagement in care among PLHIV who may feel well or face other psychosocial, structural, and health system barriers.^10-12^There is conflicting evidence regarding the acceptability and appropriateness of immediate ART initiation. While Sharma et al (2015) and Katirayi et al (2016) demonstrated > 90% acceptance rates for immediate ART, Katirayi and colleagues (albeit among pregnant women) reported that issues such as denial, shock, life-long treatment, or the requirement for partner consultation hampered uptake and adherence to rapid ART initiation.^13-14^ Furthermore, randomized controlled studies conducted in Zambia and Malawi revealed that at treatment initiation individual factors (e.g., fear of lifetime commitments and side effects, denial, substance abuse), household factors (e.g., power relations, livelihood demands), and health system factors (e.g., distance to the clinic, congestion, poor record-keeping, health care workers attitude) undermined engagement in care.^15,16^ These factors, coupled with traditional approaches to the delivery of health services, led to attrition of >30% early loss within one month of initiation.^15-17^

Current evidence on interventions that promote engagement of PLHIV following rapid ART initiation is limited.^5,13,18^ Significant questions persist about the acceptability of the universal test and treat (UTT) strategy and how it affects newly diagnosed PLHIV regarding engagement in HIV care and the factors that may affect engagement in routine settings.^4^ To address this research gap, we analysed routine programme data of PLHIV newly diagnosed and initiated on ART in community and health facility settings. Using the definitions offered by Grimsrud et al. (2021), we estimated the proportion of immediate engagement (i.e., newly diagnosed PLHIV who continue with ART 14 days after same-day ART initiation) and early engagement (i.e., newly diagnosed PLHIV who continue within the first six months of same-day ART initiation) among newly diagnosed PLHIV.^3^ Additionally we examined factors independently associated with immediate and early engagement in care among newly diagnosed PLHIV offered same-day ART initiation to provide insights that may inform design of program interventions that optimize UTT.

## Methods

### Study design and participants

This was a retrospective cohort study of newly diagnosed PLHIV who were offered same-day ART initiation during community-based or facility-based HTS. We included all newly diagnosed PLHIV individuals, aged 18 years or older and initiated on ART between October 2018 and January 2019 at all thirty-four MoH ART facilities that offered comprehensive HIV services including HIV Testing Services (HTS) in Lusaka District. Lusaka, the capital city of Zambia, is the most inhabited city in the country and home to approximately 3.5 million people.^19^ While national HIV prevalence among adults aged 15-59 years is 12.0%, prevalence in urban Lusaka district is higher at 15.7%.^2^ In February 2020 we extracted health facility data on registrations, HIV testing, new HIV-positive diagnoses, HIV-positive individuals already on ART, and date of ART initiation from registers and the national electronic medical record (EMR) database. To reliably determine the site of ART initiation, for every newly diagnosed PLHIV initiated on ART during the period of interest, we physically searched for corresponding evidence in the enrolment registers using the ART number (and matching age for confirmation).

### Outcomes

The outcomes were immediate engagement at 14 days and early engagement at 6 months. Immediate engagement was defined as a newly diagnosed PLHIV who visited the health facility at least once within 14 days after same-day ART initiation, whereas early engagement was defined as a newly diagnosed PLHIV who was found active (collected a drug refill) in the EMR 6 months after same-day ART initiation.

### Statistical Analysis

We summarized baseline characteristics (age and gender) using frequencies and proportions. Health facility size was categorized as number of active PLHIV; (large (>10,000 PLHIV), medium (5,000-10,000 PLHIV), small (1,000 - 5,000 PLHIV), and very small (<1,000 PLHIV). The differences in proportion by the site of initiation were assessed using the chi-squared test. We estimated immediate engagement by dividing number of newly diagnosed PLHIV who visited the health facility at least once within 14 days of same-day ART initiation by total number of newly diagnosed PLHIV and early engagement by dividing number of newly diagnosed PLHIV found active in EMR 6 months after same-day ART initiation by total number of newly diagnosed PLHIV. Pearson’s chi-squared test was used to assess association of the outcomes with key background characteristics. Poisson_Jregression with robust standard errors was used to_Jdetermine_Jfactors independently associated with immediate and early engagement in HIV care at 14 days and 6 months respectively. Statistical significance was set at p-value <0.05. In a secondary analysis, the Kaplan-Meier method was utilized to estimate the time to immediate engagement and compared time to immediate engagement between the site of initiation using the log-rank test. All analyses were performed using Stata 17 MP8 (StataCorp, College Station, TX, USA).

### Ethical Consideration

Approval to review existing, deidentified, routinely collected programmatic data was provided by the U.S. Centers for Disease Control & Prevention (2018-381), University of Zambia Biomedical Research Ethics Committee (011-12-17), University of North Carolina at Chapel Hill, USA (18-0854) and the Institutional Review Board at Washington University, St. Louis, USA (2019-11143).

## Results

### Characteristics of the study population at the time of ART initiation

A total of 12,777 newly diagnosed PLHIV from 34 Urban Health facilities in Lusaka District who were provided same day ART between October 2018 and January 2019 were included, of whom 7943 (62%) were initiated into HIV care from the outside the health facility “community” and 4834 (38%) from the health facility (**Table 1**).

**Table 1:** Baseline characteristics of the study population at the time of ART initiation.

|  | n(%) of Total | Facility Initiated<br>n(%) | Community Initiated<br>n(%) | p-value |
| --- | --- | --- | --- | --- |
| Age at ART start |  |  |  |  |
| 18-24 years | 2142(17) | 686 (14) | 1456 (18) | <0.001 |
| 25-35 years | 5814 (46) | 1607 (33) | 4207 (53) |  |
| 36-49 years | 4061 (32) | 1981 (41) | 2080 (26) |  |
| 50+ years | 760 (6) | 560 (12) | 200 (3) |  |
| **Sex** |  |  |  |  |
| Female | 7756 (61) | 2801 (58) | 4955 (62) | <0.001 |
| Male | 5021 (39) | 2033 (42) | 2988 (38) |  |
| **Facility size** |  |  |  |  |
| 1000-5000 | 2158(17) | 835 (17) | 1323 (17) | 0.76 |
| 5000-10000 | 5187(41) | 1942 (40) | 3245 (41) |  |
| <1000 | 291(2) | 107 (2) | 184 (2) |  |
| >10,000 | 5141(40) | 1950 (40) | 3191 (40) |  |
| **Total** | 12777 (100.0) | 4834(100) | 7943(100) |  |

1. Pearson's chi square test

Overall, the median age at ART initiation was 32 years (IQR 27-39) and 36 years (IQR 28-45) and 31 years (IQR 26-36) when disaggregated by facility and community respectively. More females initiated on ART in the community (62%) and at the facility (58%). Among the newly diagnosed PLHIV initiated in the community, a significant majority were between 25-35 years old (53%), compared to 33% in the facility, followed by those aged between 36-49 years old (26%), compared to 41% in the facility. In both the facility and community the older individuals 50+ years initiated the least.

### Immediate and early engagement at 14 days and 6 months of same-day ART initiation

Overall, 49% (n = 6257) immediately engaged in care within 14 days of testing and same-day ART initiation with no differences in proportions of engagement by facility (49%) and community (49%) initiation (**Table 2**).

**Table 2:** Proportion of Immediate and Early Engagement at 14 days and 6 months of same day ART initiation.

| Background characteristics | n(%) of Total<br>N= | Immediate engagement n (%) | p-value | Early Engagement n(%) | p-value |
| --- | --- | --- | --- | --- | --- |
| **Age at ART start** |  |  |  |  |  |
| 18-24 years | 2142 (17) | 984(46) | <0.001 | 1043(47) | <0.001 |
| 25-35 years | 5814 (46) | 2796(48) |  | 3304(57) |  |
| 36-49 years | 4061 (32) | 2100(52) |  | 2560(63) |  |
| 50+ years | 760 (6) | 377(50) |  | 452(63) |  |
| **Sex** |  |  |  |  |  |
| Female | 7756 (61) | 3730(48) | 0.013 | 4348(56) | <0.001 |
| Male | 5021 (39) | 2527(50) |  | 3011(60) |  |
| **Site of Initiation** |  |  |  |  |  |
| Facility | 4834 (38) | 2386(49) | 0.494 | 2875(59) | 0.001 |
| Community | 7943 (62) | 3871(48) |  | 4484(56) |  |
| **Facility Size** |  |  |  |  |  |
| 1000-5000 | 2158(17) | 1084(50) | <0.001 | 1420(66) | <0.001 |
| 5000-10000 | 5187(41) | 2249(43) |  | 3115(60) |  |
| <1000 | 291(2) | 142(49) |  | 160(55) |  |
| >10,000 | 5141(40) | 2782(54) |  | 2664(52) |  |
| **Total** | 12777 (100.0) | 6257(49) |  | 7359(58) |  |

Immediate engagers were male (p=0.013), aged 36-49 years old (p<0.001), and received ART services from large health facilities (p<0.001). At 6 months, overall engagement increased to 58% with strong evidence of differences by age, sex, site of initiation and health facility size. Early engagement increased with age, with the lowest observed among adults aged 18-24 years (49%) and highest in the older adults 50+ years old (63%; (p<0.001)). Males were significantly more engaged at 6 months (60%; p<0.001) whilst PLHIV who received ART services from large health facilities had the lowest early engagement rates (52%; p<0.001). (Table 2).

### Factors associated with Immediate and Early Engagement

In the Poisson regression, immediate engagement was independently associated with age, with those between 36-49 years (aRR 1.12; 95% CI 1.06-1.18) most likely to be engaged and the youngest age group (18-25 years of age) least likely to be immediately engaged (**Table 3**).

**Table 3:** Factors associated with immediate and early engagement among newly diagnosed HIV patients.

|  | Immediate Engagement |  |  |  | Early Engagement |  |  |  |
| --- | --- | --- | --- | --- | --- | --- | --- | --- |
|  | RR (95% CI) | p-value | ARR (95% CI) | p-value | RR (95%CI) | p-value | ARR (95%CI) | p-value |
| Age |  |  |  |  |  |  |  |  |
| 18-24 years | Ref |  | Ref |  | Ref |  | Ref |  |
| 25-35 years | 1.05(0.99-1.1) | <0.001 | 1.05(0.99-1.1) | <0.001 | 1.17(1.11-1.23) | <0.001 | 1.16(1.1-1.22) | <0.001 |
| 36-49 years | 1.13(1.07-1.19) |  | 1.12(1.06-1.18) |  | 1.29(1.23-1.36) |  | 1.27 (1.21-1.34) |  |
| 50+ years | 1.08(0.99-1.18) |  | 1.07(0.98-1.16) |  | 1.22(1.14-1.31) |  | 1.20 (1.11-1.29) |  |
| Sex |  |  |  |  |  |  |  |  |
| Female | Ref |  | Ref |  | Ref |  | Ref |  |
| **Male** | 1.05(1.01-1.08) | 0.013 | 1.03(0.99-1.07) | 0.135 | 1.07(1.04-1.1) | <0.001 | 0.93(0.86-1) | 0.06 |
| **Site of ART Initiation** |  |  |  |  |  |  |  |  |
| **Facility** | **Ref** | 0.493 | **Ref** |  | **Ref** |  | **Ref** |  |
| **Community** | 0.99(0.95-1.02) |  | 1(0.97-1.04) | 0.848 | 0.95(0.92-0.98) | 0.001 | 0.97(0.94-1) | 0.088 |
| **Facility size** |  |  |  |  |  |  |  |  |
| 1000-5000 | **Ref** |  | **Ref** |  | **Ref** |  | **Ref** |  |
| 5000-10000 | 0.86(0.82-0.91) | <0.001 | 0.86(0.82-0.91) | <0.001 | 0.91(0.88-0.95) | <0.001 | 0.91(0.88-0.95) | <0.001 |
| <1000 | 0.97(0.86-1.1) |  | 0.97(0.86-1.1) |  | 0.84(0.75-0.93) |  | 0.83(0.75-0.93) |  |
| >10000 | 1.08(1.03-1.13) |  | 1.08(1.03-1.13) |  | 0.79(0.76-0.82) |  | 0.79(0.76-0.82) |  |

The pattern of immediate engagement varied by health facility size, with individuals accessing ART services from large facilities (>10,000PLHIV) more likely to engage (aRR 1.08; 95% CI: 1.03-1.13; p-value <0.001) and those from health facilities (1,000-5,000 PLHIV) least likely to engage. There were no differences of immediate engagement by gender or site of ART initiation. At 6 months, newly diagnosed PLHIV engaged in the ART program were more likely to be older than 35-49 years (aRR 1.27; 95% CI 1.21-1.34; p<0.001). The younger age group (18-24 years) and PLHIV receiving care from large facilities (RR 0.79; 95% CI 0.76-0.82; p <0.001) were less likely to be engaged.

For most individuals, time from ART initiation to immediate engagement was short, 15 days median time (IQR 13-37). Survival patterns were similar with no differences in time to immediate engagement between newly diagnosed PLHIV who initiated from the facility compared to those initiated from the community (p>0.05) (Fig 1)

**Figure 1:**
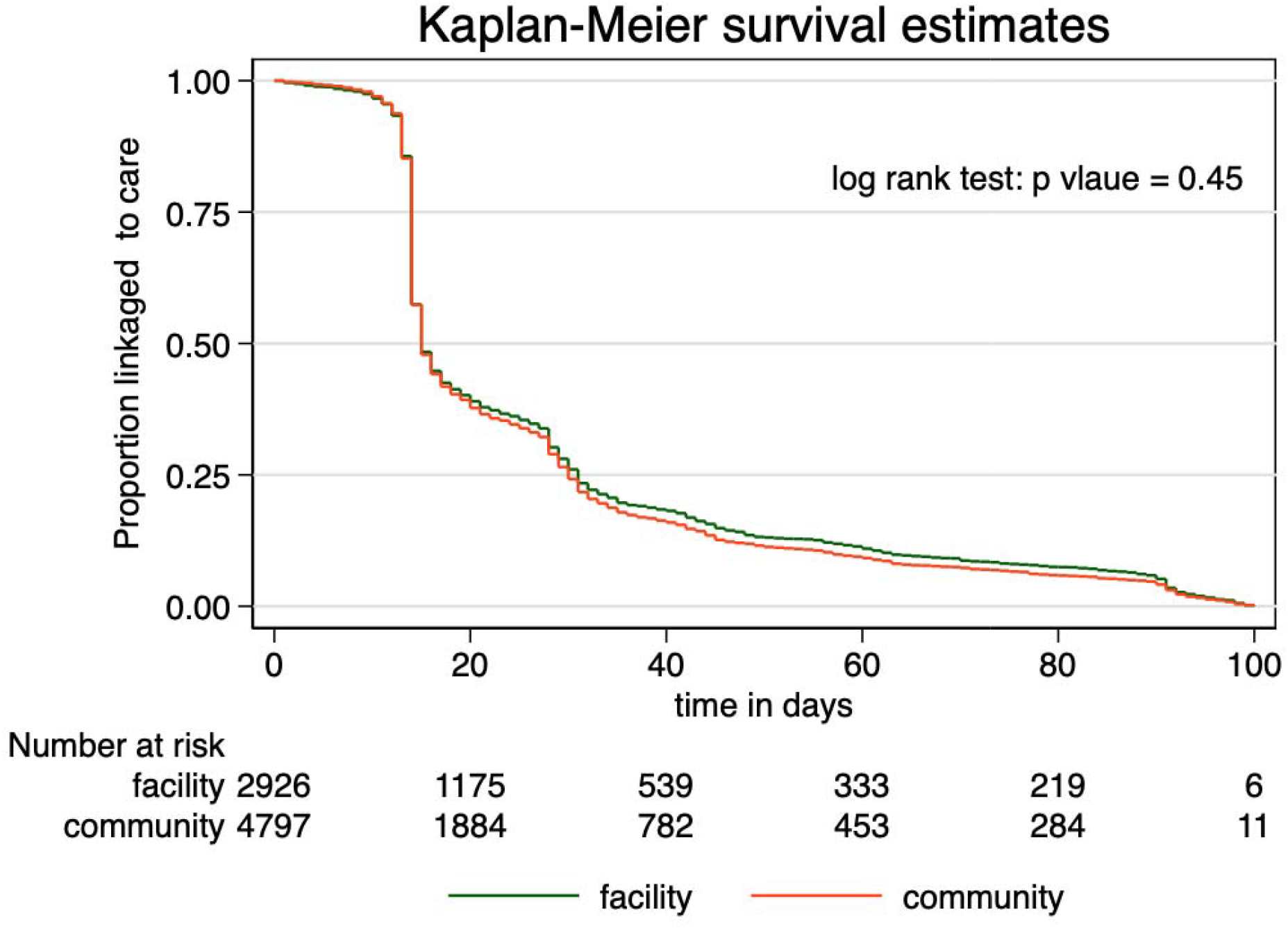
Kaplan Meier Curve Time to First Visit after ART initiation in the first 100 days

## Discussion

The study showed that out of facility “community” same day test and treat reached a higher proportion of PLHIV (62%) than facility same-day test and treat with non-significant differences in the engagement at 14 days and 6 months by the site of initiation. These study findings align with the CASCADE trial, which promotes out-of-facility testing and treatment initiation to optimally address the first and second 95 treatment targets.^20^ We found that <50% of PLHIV initiated on the same day ART immediately engaged within 14 days (49% in both community and facility) whilst at 6 months, 57% engaged in community and 56% facility respectively). This gradual increase in engagement over time is supported by findings from Grimsrud et al. (2020) and Rivka R et al. (2020) who acknowledged that newly diagnosed PLHIV require time and continued support to accept their diagnosis following testing and treatment initiation.^3,17^

Few studies have reported on immediate and early engagement after same-day ART initiation in in the community. Overall, our findings on immediate and early engagement in Lusaka compare well with a Nigerian study that reported that 51% of newly diagnosed adult PLHIV were lost to follow-up (LTFU) within the first 30 days following ART initiation compared to 30% among the previously diagnosed PLHIV (p<0.0001).^21^ Furthermore, a South African study reported significantly higher program losses at six months among same-day PLHI initiators compared to PLHIV initiating ART 1 day after HIV diagnosis (30.1% vs 21.4%).^17^ However, in both studies, PLHIV were initiated on same-day ART within health facilities.

We report that only 58% of newly diagnosed PLHIV were engaged within 6 months of ART initiation contrary to findings of improved engagement and retention after home-based testing and ART initiation reported in other studies.^22-26^ However, these studies that demonstrated significant improvement in engagement and retention after same day testing and ART initiation in the community provided incentives and nudges such as transport vouchers, telephone messages, and tracing of PLHIV who did not link to care.^22-26^Therefore, the lower engagement we found in routine settings amplifies that significant efforts are required to adequately address the unique requirements of newly diagnosed PLHIV.^27^

The finding of no difference in immediate engagement by the site of ART initiation (“community” vs health facility) runs counter to Labhart et al.’s randomized controlled trial (RCT), which showed that adult PLHIV initiated on ART in the community in rural Lesotho were significantly linked to care and engaged at 3 months when compared to those initiated at health facilities (68.6% (94) versus 43.1% (59)).^20^ The contrast between the two studies may be explained by differences in the local context, clinical procedures, and study methodologies that may affect linkage and engagement. The Labhart et al study randomized participants whilst this was an observational cohort study, thus subject to selection bias, as individuals enrolled in the community may be different from those enrolled in the health facility. Furthermore, Labhart et al conducted the study in a rural setting, sampled a small number of individuals (278), and used trained nurses to provide ART initiation at home. Whereas this study was conducted in an urban setting with ART initiation in the community provided by trained Lay Health Personnel. Additional research is therefore necessary to understand the substantial heterogenicity of linkage and engagement in care in different settings.

Our study supports the recommendations of providing targeted interventions that support engagement along the HIV care cascade.^3^ We found that immediate engagement varied among the age groups, with the 18-24 years old delaying while the 36-49 years old engaged in care quicker. These findings are corroborated by the PopART study that observed differences in linkage into care by age groups with older adults >45 years linked earlier than those aged 15-25 years old.^29^ Furthermore our study showed significant differences in 6-month early engagement by health facilities size, with attrition highest in large facilities with PLHIV load > 10,000 (aRR 0.79; 95% CI 0.76-0.82). The East African (EA)-IeDEA Consortium’s retrospective cohort study in 29 affiliated sites equally showed that clinical burden was associated with increased loss to follow-up among those who engaged with care thus underscoring the effects of high patient load on successful engagement of PLHIV.^29^ Considering the health system pitfalls in public health facilities and rapid scale-up of ART in the country, this study supports guidance that health facility level factors of clinical burden may be addressed to a certain extent by differentiated service delivery (DSD) models tailored for newly initiated PLHIV.^30^

Our study has various strengths that should be recognized, we analysed programmatic data thus provided an opportunity of describing immediate and early engagement of newly initiated PLHIV initiated on the same day ART in real-life settings thereby adding valuable insights to “community” HTS and ART initiation as well as supplement the knowledge provided by the CASCADE RCT.^20^ Furthermore, the utilization of routine data from multiple health facilities in urban Lusaka resulted in a large sample size that is likely to be representative of PLHIV who initiated ART in urban health facilities in the country although may be extrapolated with caution to many urban setting in SSA. The study however had some limitations, data only included PLHIV that had started ART during October 2018 -January 2019, this short period increased possibility of biases due to temporal factors that could not be adjust for. There is no information on individuals that did not engage, it is possible that these individuals were already on ART and did not want to disclose their status. Additionally, there was incomplete information and substantial missingness in routine data therefore, key variables such as the viral load, CD4, body mass index (BMI), WHO disease staging, education level and other sociodemographic characteristics could not be examined, thus resulted in few variables analyzed and limited adjustment of these potential confounders that may affect immediate and early engagement in HIV care. Notably, the engagement outcomes were not affected by data missingness. Lastly the study setting was urban, and findings may lack generalizability to populations in rural areas. Given these limitations, larger cohort studies with longer observation times and more attention to optimal data collection are required to confirm these results.

## Conclusion

This retrospective cohort study on same-day ART initiation of newly diagnosed PLHIV in the community reinforces the concept that immediate and early engagement may differ by gender, age, and local context. Advocating for targeted approaches in the implementation of same day ART initiation that can motivate key actors to improve routine program performance and accelerate attainment of UNAIDS targets.

## Data Availability

The Government of Zambia allows data sharing when applicable local conditions are satisfied. In this case, the data from the study will be made available to any interested researchers upon request. The CIDRZ Ethics and Compliance Committee is responsible for approving such request. To request data access, one must write to the Committee chair/Chief Scientific Officer, Dr. Roma Chilengi, or the Secretary to the Committee/Head of Research Operations, Ms. Hope Mwanyungwi mentioning the intended use for the data. The Committee will then facilitate review and authorization to release the data as requested. Data requests must include contact information, a research project title, and a description of the intended use.

## Authors’ contributions

MMM: Lead author, framed the study and wrote the first draf

OC: Conducted data analysis

EK: Supervised the data collection

GM: Assisted the underlying data processes

KS: Assisted with implementaion process.

VK: Advised implementaion details

JP: Assisted with the analysis

ML: Assisted the underlying data processes

SB: Lead advisor on data analysis

AS: Assisted with the writing

CB: Lead for implementaion process

## Acknowledgements

We would like to thank the Zambian Ministry of Health for their efforts to ensure that no one is left behind. We would also like to thank the people living with HIV and the health care workers who continue to work tirelessly to provide care to the patients.

## Copyright

This is an open access article distributed under the terms of the <u>Creative Commons Attribution License</u>, which permits unrestricted use, distribution, and reproduction in any medium, provided the original author and source are credited.

